# Inverse association between coronary artery calcification and plasma pyrophosphate: systemic deficiency and local compensation

**DOI:** 10.1101/2025.08.21.25334157

**Authors:** Martin Várhegyi, Dénes Juhász, Anna Lovrics, Emese Bata, Dániel Kovács, Bálint Szilveszter, Ádám Jermendy, Béla Merkely, Dávid Szüts, Jeremy Lagrange, Jean-Baptiste Michel, Magnus Bäck, Tamás Arányi, Anikó Ilona Nagy, Flora Szeri

## Abstract

Plasma inorganic pyrophosphate (PPi) is a potent endogenous inhibitor of vascular calcification, yet its relevance to coronary artery calcification (CAC) remains unclear, largely due to technical challenges in measurement that have limited clinical investigation. Here, we examined associations between circulating PPi and CAC in a prospective cohort of 150 cardiovascular patients. PPi levels were positively associated with serum phosphate, body mass index, and self-reported vitamin D intake, and inversely associated with alkaline phosphatase (ALP) activity and left circumflex artery calcification (LCx CAC) across the entire cohort. In patients with established calcification, LCx CAC showed an inverse association with both PPi and ALP activity, consistent with a potential local compensatory modulation of the PPi–ALP–phosphate axis. Ex vivo and in vitro analyses further supported this concept, demonstrating increased PPi release from atherosclerotic coronary arteries and calcifying vascular smooth muscle cells.

These findings provide the first clinical evidence linking reduced circulating PPi with increased CAC and support the possibility of a dual mechanism, whereby systemic PPi deficiency may promote coronary calcification, while diseased vessels may locally upregulate PPi to limit further mineral deposition. Together, these results position PPi as a potential biomarker and therapeutic target in coronary artery disease.

**GRAPHICAL ABSTRACT:** 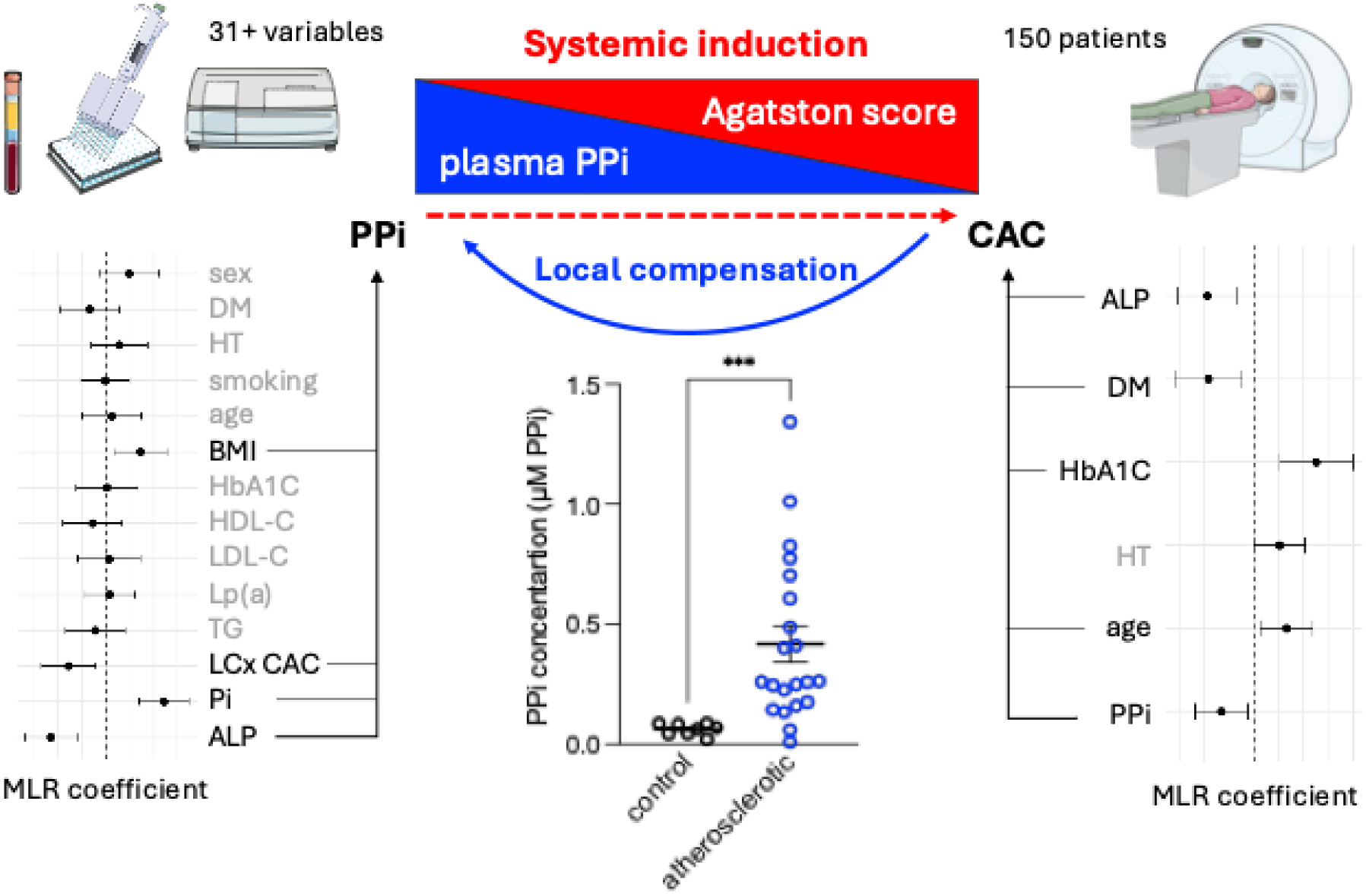

## INTRODUCTION

Coronary artery calcification (CAC) is strongly associated with coronary artery disease (CAD) and cardiovascular disease, a leading cause of death worldwide (1–4). In asymptomatic patients, the presence and extent of CAC reflect the cumulative burden of subclinical atherosclerosis and provide valuable prognostic information beyond traditional risk factors (5–7). As an independent predictor of adverse cardiovascular outcomes including myocardial infarction, stroke and cardiovascular mortality (8, 9), CAC becomes increasingly prevalent with advancing age. The two distinct types of CAC are not distinguishable with cardiac computed tomography angiography (CCTA): the frequent atherosclerotic calcification primarily affects the tunica intima, while the less prevalent medial calcification (also known as Monckeberg’s arteriosclerosis) takes place in the tunica media. The main mechanism of intimal CAC includes endothelial dysfunction, lipid accumulation and inflammation, plaque formation, vascular smooth muscle cell (VSMCs) activation, osteogenic differentiation, and calcium deposition (10). However, in medial CAC, the key mechanism is the osteochondrogenic transdifferentiation of VSMCs, with subsequent matrix vesicle release and extracellular matrix remodeling, arterial wall stiffening leading to hypertension, cardiac hypertrophy, and heart failure (11). The etiology of CAC is multifactorial (12), encompassing genetic predisposition, environmental exposures, and modifiable lifestyle factors. Despite ongoing research, the underlying molecular mechanisms remain incompletely elucidated, and targeted therapeutic strategies are currently lacking (13). The established CAC risk factors include aging, male sex or post menopause, dyslipidemia, hypertension, obesity and smoking (3, 14). Additionally, in hemodialysis and advanced chronic kidney disease, especially the medial type of CAC is significantly increased, accompanied by hyperphosphatemia, elevated parathyroid hormone (PTH) and increased fibroblast growth factor-23 (FGF-23) levels (15). Moreover, metabolic syndrome and diabetes accelerate medial type CAC (16, 17) through the combined effects of endothelial dysfunction, inflammation, altered calcium-phosphate balance, hyperglycemia, and oxidative stress (18, 19). Sedentary lifestyle and western-type diet further contribute to CAC progression. Finally, ethnicity (20), and genetic predisposition to elevated lipoprotein(a) [Lp(a)] levels, along with insufficient levels of calcification inhibitors, such as matrix Gla protein (MGP) (21) and deficiency of ectonucleotide pyrophosphatase/phosphodiesterase 1 (ENPP1) (22–24) may drive or contribute to the acceleration of CAC.

ENPP1 is the only extracellular enzyme that can generate inorganic pyrophosphate (PPi) (25), a key endogenous inhibitor of vascular calcification. The predominantly hepatocyte-specific transporter ABCC6 and the more ubiquitously expressed ANKH protein mediate the release of ATP into the circulation (26, 27) supplying the substrate for ENPP1 to generate PPi. Tissue-nonspecific alkaline phosphatase (TNAP) subsequently degrades PPi into inorganic phosphate (Pi), thereby removing a critical endogenous inhibitor of calcification (28). Consequently, hereditary deficiencies in factors responsible for PPi generation result in absent or reduced systemic PPi levels and are causative of rare calcification disorders; Generalized Arterial Calcification of Infancy (GACI) (23), Pseudoxanthoma Elasticum (PXE) (26), and Craniometaphyseal Dysplasia (CMD) (29). Fully or incompletely penetrant loss of function ABCC6 variants cause PXE (30), in which cardiovascular manifestations have been reported (31–33). Interestingly, the most frequent ABCC6 nonsense variant has been associated with an increased CAD prevalence (34). Importantly, PPi levels are significantly decreased in end-stage chronic kidney disease patients and in hemodialysis, coinciding with progressed medial calcification, and increased alkaline phosphatase (ALP) activity (35, 36). A recent study directly linked PPi deficiency to diabetes in animal models, demonstrating that elevated glucose levels lead to downregulation of ENPP1 and upregulation of ALP, thereby increasing the propensity for vascular calcification (37). Furthermore, we have recently shown that aortic valve calcification, after adjustment for traditional risk factors, positively associates with increased plasma Pi/PPi ratio in a heterogenous cohort of cardiovascular patients (38).

Based on this accumulating evidence, we hypothesized that circulating inorganic pyrophosphate (PPi) may contribute to the pathogenesis of coronary artery calcification. Here, we provide the first systematic evaluation of clinical and biochemical correlates of plasma PPi in relation to CAC and explore whether PPi-generating local compensatory mechanisms may operate within the coronary vasculature using ex vivo human coronary artery explants and in vitro vascular calcification models.

## METHODS

### Patient population

Caucasian patients referred for clinically indicated CCTA were prospectively enrolled in this study. Patients were contacted before enrolment to confirm their willingness to participate and to ensure adherence to the pre-procedural requirements crucial for proper determination of plasma PPi level (see requirements at blood sampling). Patients with conditions that may interfere with either CAC quantification or plasma PPi measurements were not enrolled in the study. Specifically, individuals with mineral metabolism disorders, inflammatory or autoimmune diseases or severe renal impairment (defined as an estimated glomerular filtration rate [eGFR] <30 mL/min/1.73 m², by the CKD-EPI creatinine equation) in previous medical history were not contacted, due to the known association between end-stage kidney disease and increased vascular calcification. Additionally, patients with coronary artery stents, which can impair the accuracy of CAC scoring, and individuals receiving bisphosphonate or vitamin K antagonist (e.g., warfarin) therapy, which may alter PPi metabolism and ectopic mineralization, were not enrolled. Hypertension (HT) and diabetes mellitus (DM) were highly prevalent in our patient population; therefore, we could not include these as exclusion criteria. Demographic and clinical data were obtained at the time of enrolment using a structured questionnaire, including comorbidities (e.g., DM and HT) and treatments (e.g., statins and anticoagulants). Electronic medical records were reviewed to supplement or verify the information provided. The study was approved by the Scientific and Research Ethics Committee of the Hungarian National Medical Scientific Council (BMEÜ/3473-1/2022/EKU). The study protocol complied with the declaration of Helsinki. All participants provided written informed consent for CT imaging, blood sampling, and statistical data analysis.

### Blood sampling

Patients were asked to strictly adhere to the pre-procedural requirements, including a minimum of 8 hours of fasting, abstinence from beverages other than tap water, and avoidance of strenuous physical activity prior to the examination. Venous blood samples were collected prior to the CCTA procedure using a 21-gauge (or larger) intravenous cannula, with care taken to minimize tissue trauma. If the initial venipuncture attempt was unsuccessful, a second attempt was made in the contralateral arm. Patients requiring more than two attempts were excluded from the study to avoid confounding effects from tissue injury. For plasma inorganic pyrophosphate (PPi) measurement, blood was always collected into the second vacuum tube containing K₃EDTA and gently inverted four times immediately after blood collection. To minimize pre-analytical variability, samples designated for PPi analysis were handled at room temperature with particular care, as cold or mechanical stress can significantly alter PPi concentrations. All samples were transferred to the preparative laboratory within 30 minutes of collection.

### Sample preparation and determination of routine clinical laboratory parameters

Whole blood, EDTA-anticoagulated plasma, and serum samples were used for routine clinical laboratory analyses. Whole blood was used for hematology (e.g., blood count), EDTA plasma for HbA1c (hemoglobin A1c) measurement, and serum—either from plain or gel-separation tubes—for blood chemistry and lipoprotein(a) [Lp(a)] analysis. Plasma and serum fractions were separated according to standard protocols, and all parameters were measured following the routine procedures of the Heart and Vascular Centre, Semmelweis University, Budapest, Hungary.

### Sample preparation for inorganic pyrophosphate determination

The plasma fraction from the blood harvested to a vacuum tube containing K₃EDTA (39) was isolated via two consecutive repeated centrifugations (1000g for 5 minutes at room temperature) and transferred into platelet separation tubes (Vivaspin Filtrate, 300kDa, 13279-E, Sartorius, Göttingen, Germany). The platelet-free plasma was obtained by further centrifugation at 2000 g for 30 minutes at room temperature and stored at −80°C until PPi measurement (conducted within 3-months).

### Inorganic pyrophosphate determination

The PPi content of the plasma was determined enzymatically, using the ATPS bioluminescent method, with slight modifications. First, PPi was converted into ATP in an assay containing 80 µM MgCl2, 50 mM HEPES pH 7.4, 32 mU/ml ATP Sulfurylase (M1017B, New England Biolabs, Ipswich, MA, USA) and 16 µM adenosine 5’-phosphosulfate (A5508, Sigma-Aldrich, Saint Louis, MO, USA). Samples were incubated at 37°C for 30 minutes, followed by enzyme inactivation at 90°C for 10 minutes. In the next step, ATP levels were evaluated utilizing the BacTiterGlo (Promega, Madison, WI, USA) bioluminescent assay, prepared according to the manufacturer’s instructions, and analyzed in a multimode plate reader (PerkinElmer). Plasma PPi concentrations were calculated using calibration standards and correction for initial plasma ATP concentrations (26). Plasma samples with ATP concentrations exceeding 0.6 µM were excluded from PPi analysis, based on a predefined quality control threshold to ensure appropriate blood collection and pre-analytical processing.

### Cardiac CT angiography (CCTA) acquisition and calcium scoring

CCTA scans were conducted using either a dedicated cardiac CT system (CardioGraphe, GE Healthcare, Chicago, IL, USA) or a third-generation dual-source CT scanner (SOMATOM Force, Siemens Healthineers, Forchheim, Germany), employing a prospectively ECG-triggered axial scan mode. All examinations were performed at a tube voltage of 120 kVp, with tube current modulation based on individual patient body size. For CAC assessment, non-contrast-enhanced scans were obtained with 2 mm slice thickness, applying iterative reconstruction techniques. CAC scoring was carried out offline using the Agatston method (40, 41) on a dedicated analysis workstation equipped with commercially available software (HeartBeat CS, Philips Healthcare, Cleveland, OH, USA). A single trained investigator evaluated all scans, identifying coronary calcifications on axial images. Total coronary artery calcium (sumCAC) score was calculated as the sum of Agatston scores across the major coronary arteries; left main coronary artery (LM), left anterior descending artery (LAD), left circumflex artery (LCx) and right coronary artery (RCA).

### PPi determination in conditioned media from coronary artery explants

Human coronary artery walls, including healthy and diseased arteries at different stages of atherosclerotic diseases, were collected from the Inserm human CV biobank (BB-0033-00029, U 1148, X. Bichat Hospital, Paris), included in the European network BBMRI-ERIC, in accordance with the French regular and ethical rules (BioMedicine Agency convention DC2018-3141) and the principles of the declaration of Helsinki. Approval was obtained from the French Biomedical Agency (ABM, PFS09-007 & PFS17-002) and the Institutional Ethical Review board (SC09-09-66). Tissues were obtained from deceased organ donors for kidney and/or hepatic transplantation in the absence of therapeutic uses for the heart. When collected, healthy coronary arteries were macroscopically analyzed by a trained vascular surgeon to ensure that these samples were not pathological (absence of fatty streak or atherosclerosis plaques after longitudinal opening). Explants were stratified into three categories: arteries with calcified atherosclerotic lesions, arteries with atherosclerotic lesions, or control arteries without atherosclerotic lesions. Conditioned media were prepared from small pieces (about 2 mm^3^) of tissue, weighed, and incubated for 24 h in serum-free RPMI culture medium containing 1% L-glutamine, 1% penicillin, streptomycin, and amphotericin at 37°C (5% CO_2_). For standardization, the volume of medium was adjusted to sample wet weight (6 mL per gram of tissue). Conditioned media were collected, centrifuged (14000 g for 15 min at 4°C), and stored at 80°C until PPi quantification. PPi concentrations were measured as described above, with a modification involving a fivefold increase in sample volume to account for the low PPi concentrations in the samples.

### *In vitro* calcification assay using A7r5 vascular smooth muscle cells

A7r5 rat VSMCs, obtained from ATCC, were used for *in vitro* calcification studies. Cells were maintained according to the manufacturer’s protocol in Dulbecco’s Modified Eagle Medium (DMEM) supplemented with 10% fetal bovine serum (FBS) and 100 mg/L penicillin-streptomycin. For the calcification assay, 5 × 10⁴ A7r5 cells were seeded into 12-well plates and allowed to adhere and grow for 24 hours. The assay was conducted as previously described (42). Briefly, calcification was initiated by replacing the medium with either phosphate-supplemented (3 mM Pi) or standard control medium (1 mM Pi). Media were refreshed at 72 and 120 hours, and cells along with conditioned media were harvested at 168 hours for analysis. Conditioned media collected at 72, 120, and 168 hours were used for PPi quantification. Cell viability and metabolic activity were assessed at 168 hours using the PrestoBlue (Invitrogen) assay according to the manufacturer’s instructions. For long-term calcification assessment, cells were cultured for 21 days with medium changes every three days. Calcification was confirmed on day 21 by Alizarin Red staining. PPi concentrations in the culture media were determined as previously described (43).

### Data processing, statistical and regression analysis

A total of 150 patients were enrolled, post recruitment exclusions included three patients with eGFR <30 mL/min/1.73 m², one patient with multiple liver enzyme outliers, one with a plasma PPi outlier suggestive of preanalytical error, and fifteen with plasma ATP concentrations exceeding our internal quality control thresholds. Three patients with missing data were also excluded. Non-numeric values for Lp(a) (“<0.1”) and eGFR (“>90”) were replaced with random values within physiologically plausible bounds. Binary variables were coded as 0 or 1, with 0 indicating absence (or male sex) and 1 indicating presence (or female sex). Continuous variables were evaluated for normality via the Shapiro-Wilk test. Parametric variables are expressed as mean ± SEM; nonparametric variables as median [IQR]; categorical variables as counts and percentages. Spearman correlation was calculated between each pair of variables (Supplementary Figure 1). In the case of each continuous variable, potential outliers were detected based on Rosner’s generalized extreme Studentized deviate test using RosnerTest from R’s EnvStats package. While statistically, some values were flagged as outliers, they were still retained in the analysis based on medical considerations. Lastly, the variable ATP was removed, since its purpose was solely quality control for PPi determination and not model prediction. Univariate and multiple linear regressions were performed in R (glm, MASS::polr). To prevent overfitting, highly intercorrelated variables (Spearman’s ρ > 0.6; e.g., LM, LAD, RCA CAC, cholesterol, GOT, RBC, hematocrit, creatinine) were excluded, and multicollinearity was confirmed (VIF < 5). To circumvent overfitting, where indicated, the number of predictors were further restricted, and a best-fitting model was selected from combinatory alternative models based on lowest AICc value. Model fitting was followed by diagnostic analysis to verify modeling assumptions. Linearity was assessed by plotting the residuals vs the fitted values and homoscedasticity by plotting the residuals or the square root of the absolute standardized residuals vs the fitted values. Additionally, normality was assessed by the Q-Q plot of the standardized residuals and to identify observations with both high leverage and large residuals, standardized residuals were plotted vs. the leverage. Additionally, influential observations were assessed using Cook’s distance, and standardized residuals were plotted against leverage to identify observations with both high leverage and large residuals. Additionally, a stacked bar plot of predicted class probabilities was generated for each observation.

PPi levels in conditioned media from 29 coronary artery explants were compared using a two-tailed Mann–Whitney U test. For in vitro experiments, a two-way repeated-measures ANOVA was used to assess the effects of exogenous phosphate treatment and incubation time on PPi accumulation in A7r5 cell–conditioned media across three independent experiments. Sidak’s multiple-comparisons test was applied post hoc to adjust for multiple testing. Data are presented as mean ± SD, and p values are reported where applicable. A two-sided p value < 0.05 was considered statistically significant. Statistical analyses were performed using R (v4.3.3) and GraphPad Prism (v10.5.0).

## RESULTS

### Demographic, clinical, and laboratory characteristics of patients

A total of 150 cardiovascular patients were prospectively enrolled. Recruitment was restricted to individuals without disorders, conditions, or treatments expected to confound mineral metabolism or vascular calcification analyses (see Methods). Because plasma inorganic pyrophosphate (PPi) is technically challenging to quantify and not routinely measured in clinical practice, additional stringent criteria for patient inclusion, sample collection, processing, quality control, and exclusion were applied to ensure reliable PPi assessment (see Methods). Following these procedures, 127 participants were included in the final analysis, and 31 independent variables were evaluated (see Methods and Supplementary Figure 1). Based on total coronary artery calcium (SumCAC) scores, patients were categorized as non-calcified (sumCAC = 0; n = 30) or calcified (sumCAC > 0; n = 97). As expected, CAC scores were significantly elevated across all arterial beds in the calcified group, however sex distribution was unaltered. In contrast, age was strongly associated with CAC prevalence, with 54 years [IQR: 51–66] in non-calcified individuals to 73 years [63–78] in the CAC group (p < 0.001). BMI was slightly increased among calcified patients, (28.85 ± 0.48 kg/m²; p = 0.046) compared to non-calcified (26.86 ± 0.86 kg/m²). DM status significantly increased with CAC prevalence (24.7% vs 6.7%, p = 0.038), as did hypertension (71.1% vs 43.3%, p=0.008) and statin use (48.5% vs. 23.3%, p=0.020). eGFR was decreased in the calcified group (74 [62–85] vs. 95 [83–95]), p < 0.001), though values remained above the threshold for impaired kidney function. In addition, HbA1C (38.32 [36.03 - 43.53] vs. 35.58 [33.77 - 40.90]) and Lp(a) (0.14 [0.00 - 0.75] vs. 0.00 [0.00 - 0.15]) were both increased in the calcified group. Of note, plasma PPi, phosphate levels and ALP activity were similar in both calcified and non-calcified groups.

### Determinants of plasma PPi; association with phosphate, alkaline phosphatase, BMI and CAC

It is largely unknown what factors determine plasma inorganic pyrophosphate (PPi) levels in healthy individuals or patient populations, especially in the context of CAC. To assess factors that predict PPi levels in our cohort, we first applied univariate linear regression analysis with all available variables. As shown in bold in Figure 1A, we revealed a significant positive association of PPi levels with serum phosphate (Pi) concentration, body mass index (BMI) and self-reported vitamin D (Vit D) intake, while PPi levels showed a significant negative association with serum alkaline phosphatase (ALP) activity. Of note, in univariate linear regression the association of PPi with coronary artery calcium score in the left circumflex artery (LCx CAC) and the overall burden of coronary artery calcification represented by the sum coronary artery calcium score (sumCAC) showed inverse trends but did not reach statistical significance (p = 0.060, and p=0.243, respectively). The proportion of variance in PPi levels explained by each variable is shown in Figure 1B, with significant determinants highlighted in red and non-significant ones in turquoise. Notably, the LCx CAC score ranks as the first non-significant determinant. The univariate plots are shown in Supplementary Figure 2, except for Pi, BMI and ALP, along with LCx CAC and sumCAC, which are shown in Figure 1C.

**Figure 1.**
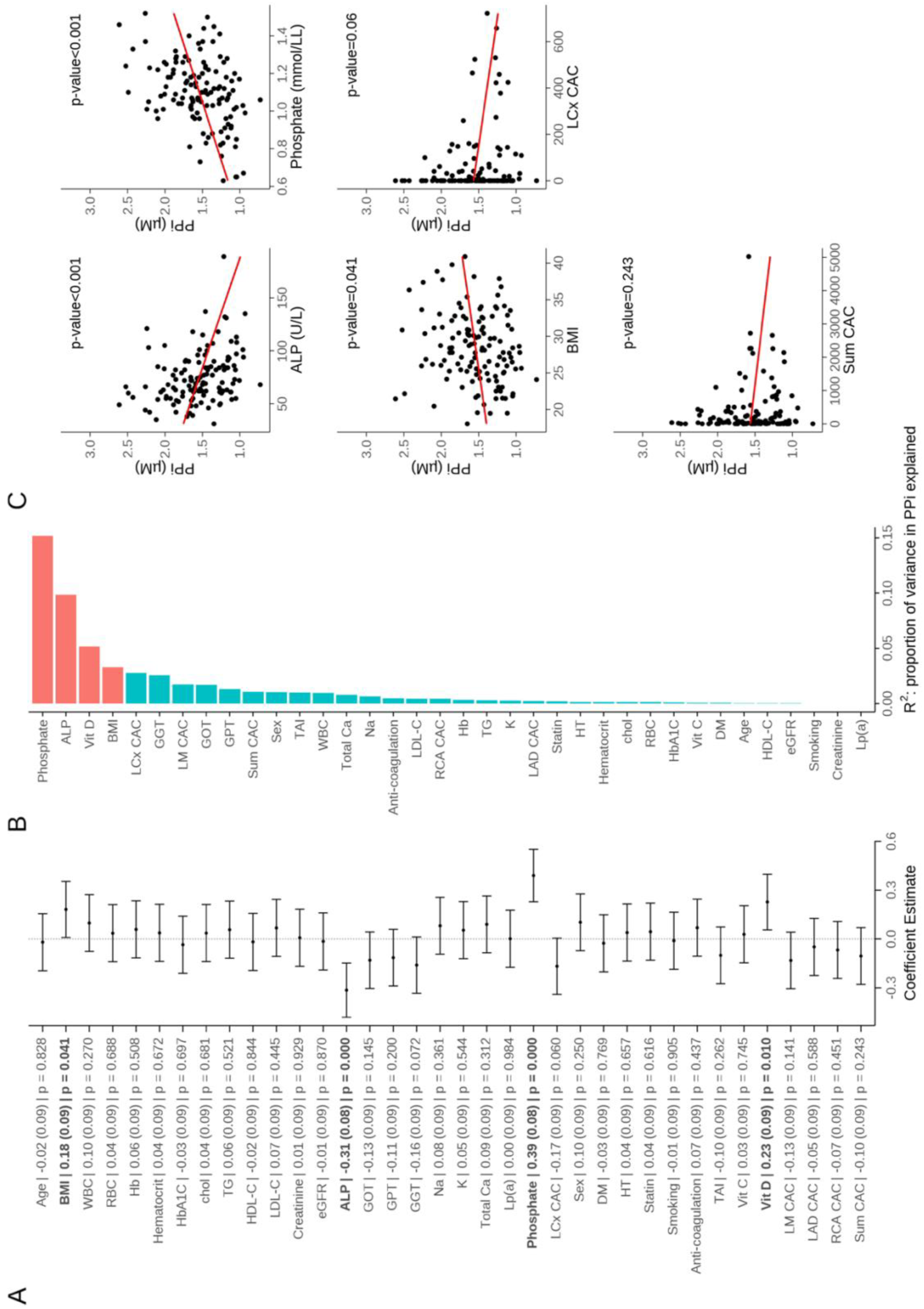
Predictors of plasma PPi level in univariate linear regression. Determinants of PPi by univariate linear regression with unselected variables. Effect size (β) with standard error and p value are indicated for each covariate (A). Proportion of variance explaining PPi levels by the variables, red indicating significant and turquoise non-significant contributors (B). Individual univariate plots depicting PPi levels in the function of ALP, Pi, BMI, LCX CAC and sumCAC (C). ALP, serum alkaline phosphatase activity; BMI, body mass index; chol, total cholesterol; DM, diabetes mellitus; eGFR, estimated glomerular filtration rate; GGT, gamma-glutamyl transferase; GOT, glutamic-oxaloacetic transaminase; GPT (U/L), glutamic pyruvic transaminase; Hb, hemoglobin; HbA1c, hemoglobin A1c; HDL-C, high density lipoprotein cholesterol; HT, hypertension; K, potassium; LAD CAC, Agatston score of the left anterior descending artery; LCx CAC, Agatston score of the left circumflex artery; LDL-C, low density lipoprotein cholesterol; LM CAC, Agatston score of the left main coronary artery; Lp(a), lipoprotein(a); Na, sodium; PPi, plasma inorganic pyrophosphate; RBC, red blood cell; RCA CAC, Agatston score of the right coronary artery; Sum CAC, Agatston score of the total coronary artery calcium; TAI, thrombocyte aggregation inhibitor; TG, triglycerides; Total Ca, total calcium; vit C, vitamin C; vit D, vitamin D; WBC, white blood cell.

To identify factors that predict the normally distributed plasma PPi levels while minimizing potential statistical bias from multicollinearity and confounding, we applied multiple linear regression (MLR) models. Consistent with previous literature, as shown in Supplementary Figure 1, we observed strong intercorrelations among the CAC scores of the four main coronary artery beds. Notably, CAS in the left circumflex artery showed the strongest inverse trend with plasma PPi level. Accordingly, we examined the predictive value of LCx CAC alongside the total coronary calcium burden for plasma PPi levels. After excluding highly correlated variables (see Methods for details), we constructed MLR models incorporating LCx CAC or sumCAC, along with variables found significant in univariate analysis (Pi, ALP and BMI, although omitting the self-reported vitamin D). To account for potential confounders, we further included established CAC risk factors: sex, age, LDL-C, HDL-C, triglycerides (TG), Lp(a), HbA1c, diabetes (DM) and hypertension (HT) status, and smoking. In MLR models containing either LCx CAC or sumCAC scores, serum phosphate (Pi), alkaline phosphatase (ALP) activity, and body mass index (BMI) consistently emerged as significant determinants of plasma PPi levels (β = 0.37, SE = 0.08, p < 0.001; β = −0.35, SE = 0.08, p < 0.001; and β = 0.22, SE = 0.09, p = 0.012, respectively; Figure 2). Notably, LCx CAC also emerged as a statistically significant predictor of plasma PPi (β = −0.24, SE = 0.09, p = 0.009; Figure 2A). By contrast, sumCAC displayed a consistent inverse association with plasma PPi, albeit with a smaller effect size that did not reach statistical significance (β = −0.14, SE = 0.09, p = 0.122; Figure 2B).

**Figure 2.**
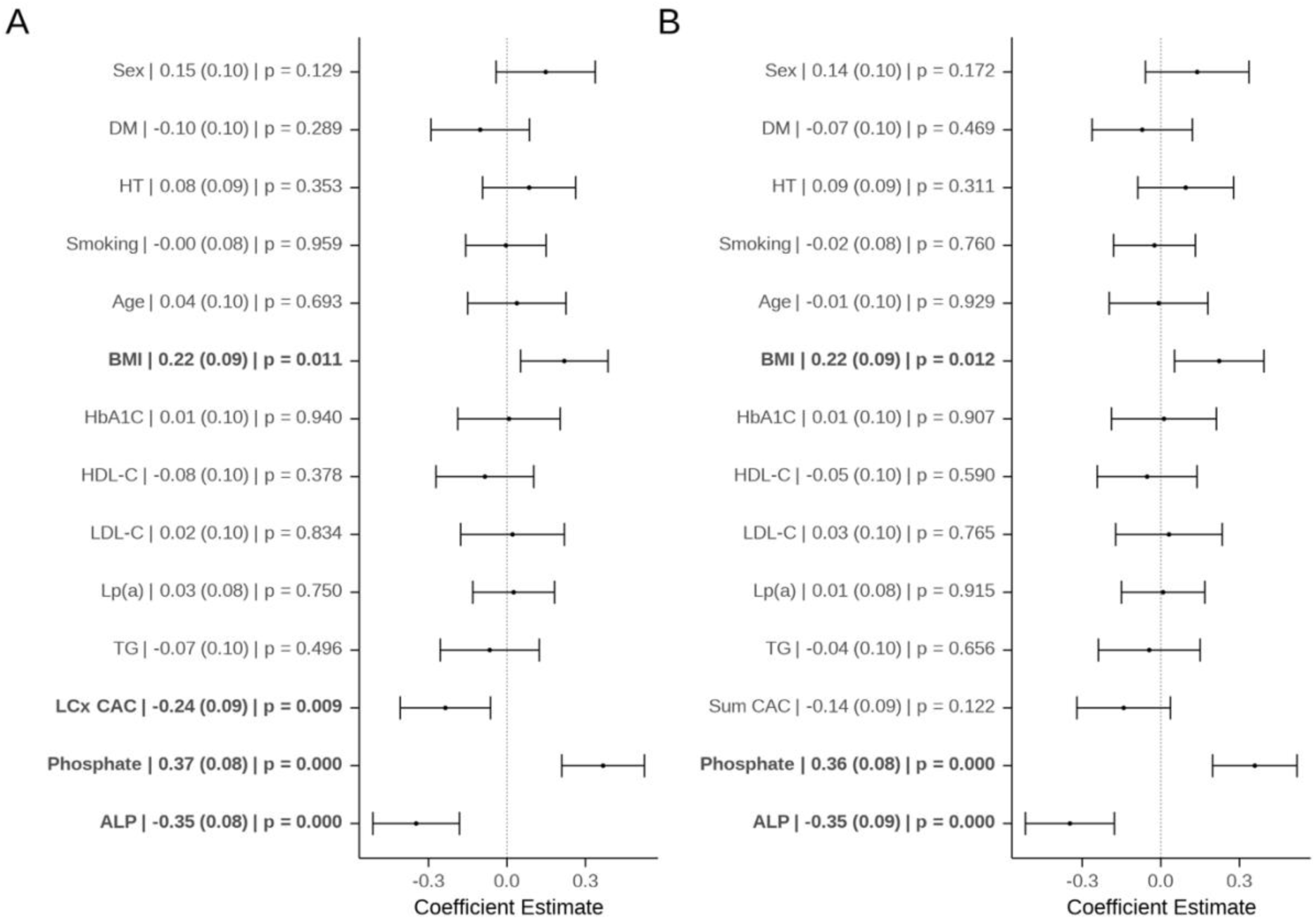
Determinants of plasma PPi level in multiple linear regression. Determinants for association of PPi with LCx CAC score and other covariates (A) or sumCAC score and additional covariates (B) in the entire cohort. Effect size (β) with standard error and p value are indicated for each covariate. ALP, serum alkaline phosphatase activity; BMI, body mass index; DM, diabetes mellitus; HbA1c, hemoglobin A1c; HDL-C, high density lipoprotein cholesterol; HT, hypertension; LCx CAC, Agatston score of the left circumflex artery; LDL-C, low density lipoprotein cholesterol; Lp(a), lipoprotein(a); Sum CAC, Agatston score of the total coronary artery calcium; TG, triglycerides.

### Determinants of CAC; association with PPi and alkaline phosphatase activity in the calcified cohort

Predictors of coronary artery calcification are well established; however, its association with plasma inorganic pyrophosphate has not previously been examined. To address this, we analyzed determinants of CAC using MLR in patients with established coronary calcification. Restricting the analysis to calcifying patients (n = 97) minimized distributional bias introduced by a substantial proportion of zero calcium scores (n = 30). In the calcified cohort, left circumflex artery (LCx) CAC was inversely associated with plasma PPi levels in MLR models adjusted for sex, age, smoking status, body mass index, diabetes mellitus (DM), hypertension, LDL-C, HDL-C, triglycerides, lipoprotein(a), HbA1c, serum phosphate, and alkaline phosphatase (ALP) activity (Figure 3A). Notably, DM also showed a significant inverse association with LCx CAC, while none of the other traditional risk factors associated with LCx CAC.

**Figure 3.**
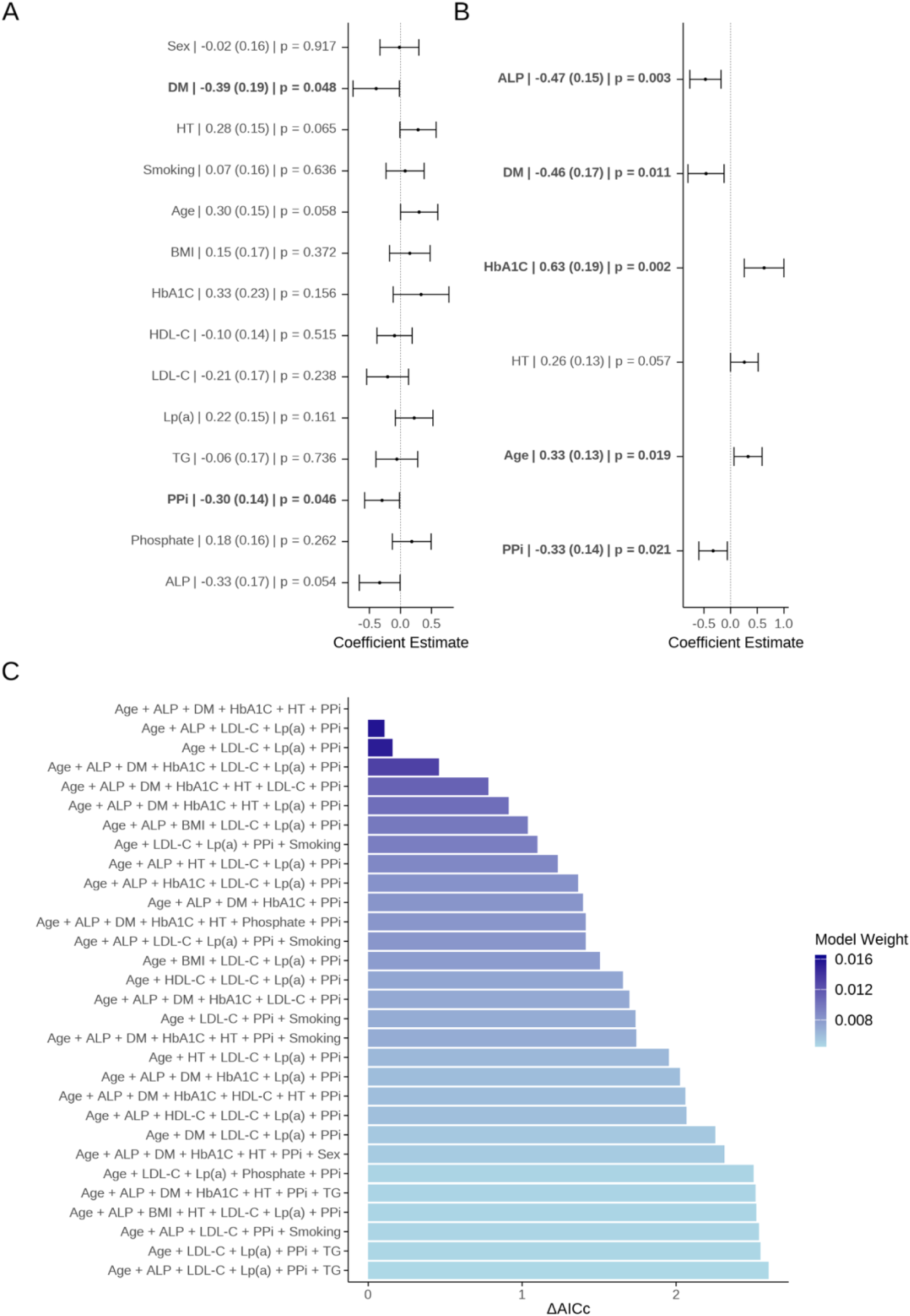
Determinants of CAC in calcifying patients in multiple linear regression (MLR). Determinants for association with LCx CAC score with adjustment for major confounders (A). Determinants for association with LCx CAC score in the best of the restricted MLR models (B). In A and B effect size (β) with standard error and p value are indicated for each covariate. Ranking of the best-fitting restricted MLR models, based on corrected Akaike information criterion (AICc) (C). The covariates included in the restricted MLR models and the models’ alteration from the best-fitting MLR model (ΔAICc) is indicated. ALP, serum alkaline phosphatase activity; BMI, body mass index; DM, diabetes mellitus; HbA1c, hemoglobin A1c; HDL-C, high density lipoprotein cholesterol; HT, hypertension; LCx CAC, Agatston score of the left circumflex artery; LDL-C, low density lipoprotein cholesterol; Lp(a), lipoprotein(a); PPi, plasma inorganic pyrophosphate; TG, triglycerides.

To reduce the risk of overfitting, we generated a series of restricted MLR models using a combinatorial approach. The maximum number of independent variables was limited to seven, selected from the fourteen variables included in the full MLR model (see above). Plasma PPi and age—given PPi’s relevance to this study and age’s well-established association with CAC—were retained in all models, while the remaining covariates were permuted. Figure 3B depicts the best restricted MLR model, whose selection was based on corrected Akaike information criterion (AICc), balancing model fit and parsimony (Figure 3C). The best-fitting model (Figure 3B) included six independent variables and confirmed a significant inverse association between LCx CAC and plasma PPi (effect size −0.33, SE = 0.14, p=0.021). Positive associations emerged for HbA1c and age (β = 0.63, SE = 0.19, p = 0.002; and β = 0.33, SE = 0.13, p = 0.019, respectively), whereas hypertension showed a positive trend that did not reach statistical significance (β = 0.26, SE = 0.13, p = 0.057). In contrast, the inverse association with diabetes mellitus was retained (β = −0.46, SE = 0.17, p = 0.011). Finally, LCx CAC exhibited a strong inverse association with alkaline phosphatase (ALP) activity (β = −0.47, SE = 0.15, p = 0.003). Consistently, sumCAC in calcifying patients exhibited a similar but non-significant inverse trend with plasma PPi and ALP (β = −0.14, SE = 0.10, p = 0.179; and β = −0.16, SE = 0.10, p = 0.108, respectively) and a positive trend with Pi (β = 0.13, SE = 0.10, p = 0.214) (Supplementary Figure 3).

### Evidence for a local compensatory PPi response

Collectively, the inverse associations of CAC with plasma PPi and ALP activity in the calcifying patients support dysregulation of the PPi–ALP axis in patients with established calcifications and are compatible with a local compensatory response to mineral stress in calcifying coronary arteries.

To investigate this hypothesis experimentally, we studied PPi accumulation in media conditioned with human coronary artery explants. We used explants from 29 donors with or without clinically confirmed atherosclerotic lesions. Comparative analysis revealed significantly higher PPi concentrations in the media from atherosclerotic arteries (n=21, 0.417 ± 0.075 µM PPi, p = 0.0002), compared to that of healthy arteries (0.0629 ± 0.0086 µM PPi) (Figure 4A).

**Figure 4.**
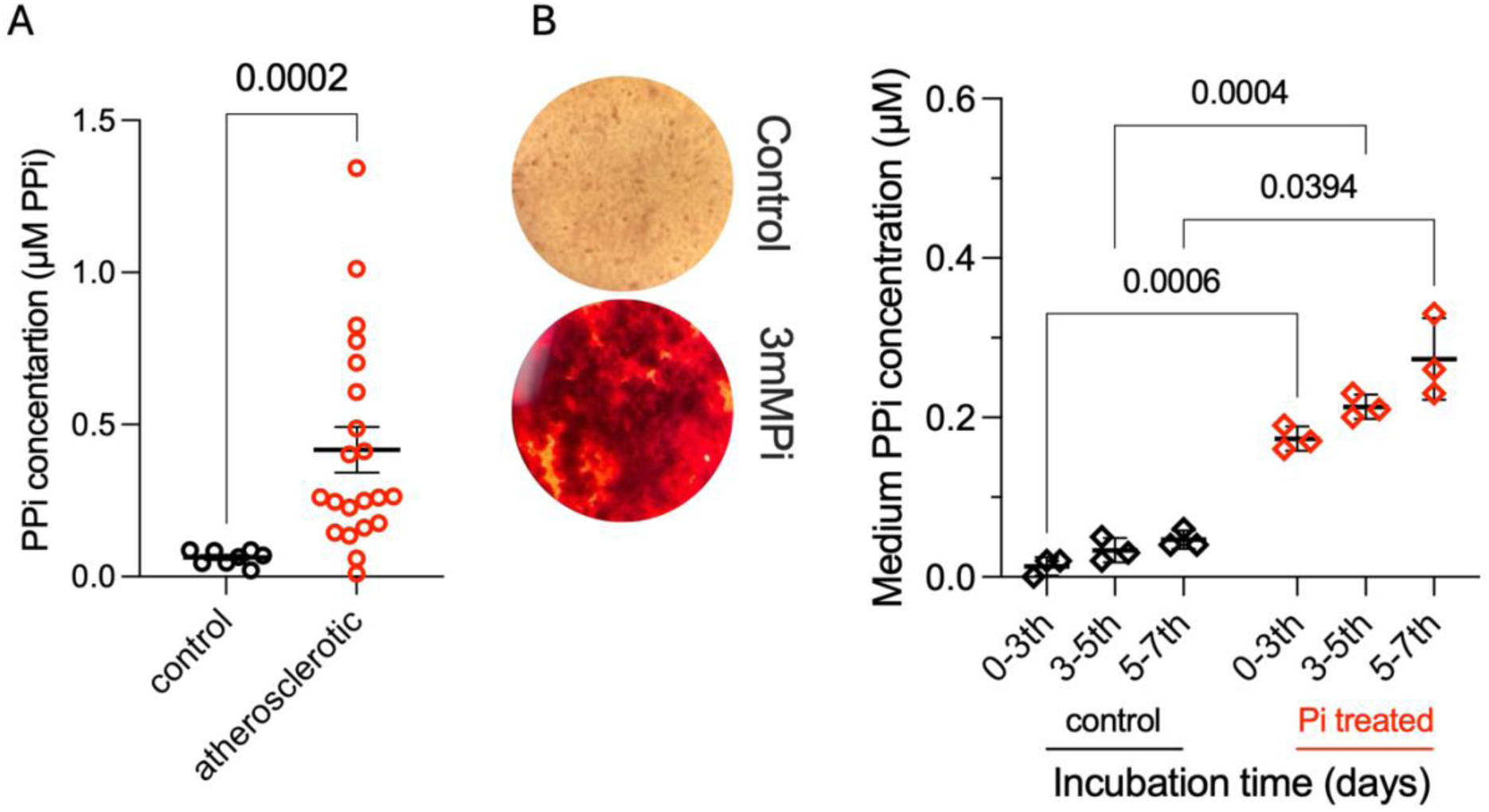
PPi level relative to pathologies or mineral stress ex-vivo and in vitro. PPi concentration of conditioned media containing coronary artery explants obtained from 29 patients (A). Explants were stratified into control arteries with no atherosclerotic lesions (n = 8, black) or arteries with atherosclerotic lesions (n = 21, red) (B). Mann-Whitney U test reveals a significant increase in PPi concentration in the conditioned media of pathologic arteries. Calcification in A7r5 rat VSMCs upon phosphate (Pi) stimulus detected by Alizarin red staining, and accumulation of PPi in the media of Pi treated and untreated A7r5 cells (n = 3 independent experiments) (B). Two-way ANOVA with Sidak’s multiple comparison post hoc test reveals phosphate and time-dependent significantly increased PPi accumulation in media of calcifying VSMC.

To corroborate this striking observation, we employed an *in vitro* model using A7r5 vascular smooth muscle cells, a widely used cell line for studying vascular calcification (44). Cells were exposed to phosphate-enriched medium (3mM Pi) to induce osteogenic differentiation and matrix calcification, confirmed by positive Alizarin Red staining (Figure 4B). Notably, Pi induced mineral stress led to a significant, time-dependent increase in extracellular PPi levels in the culture medium of calcifying A7r5 cells (0.173 ± 0.015 µM PPi, p =0.0006; 0.213 ± 0.015 µM PPi, p = 0.0004 and 0.273 ± 0.051 µM PPi, p = 0.0394) compared to cells maintained in standard 1 mM Pi containing medium (0.013 ± 0.012 µM PPi; 0.033 ± 0.015 µM PPi; 0.047 ± 0.012 µM PPi) incubated between the 0-3^th^, 3-5^th^ and 5-7^th^ days, respectively (Figure 4B).

## DISCUSSION

### Clinical relevance and challenges of plasma PPi measurement

Systemic PPi deficiency, whether due to impaired synthesis or accelerated degradation, is a recognized contributor to pathological calcification. Even in physiological states, such as healthy pregnancy, plasma PPi levels decline (45), potentially contributing to the increased cardiovascular risk observed in multiparous women (46). Despite PPi’s known role as an endogenous inhibitor of calcification (47), clinical studies examining its involvement in vascular disease are scarce. This limitation stems primarily from the analytical challenges of quantifying PPi in plasma, requiring careful patient inclusion, reliable assays and strict pre-analytical processing. Although recent assay advancements have improved PPi determination (39, 48), remaining analytical constrains and the need for rigorous pre-analytical processing continues to hinder its clinical implementation. Despite rare hereditary diseases with low PPi levels and vascular calcification (31, 49), data for PPi levels in healthy individuals and multifactorial common diseases are scarce (36, 38, 48). To date, no studies have directly examined the association between circulating PPi levels and CAC. Our study addresses this gap in a heterogenous cardiovascular patient cohort, while also identifying clinical and biochemical determinants of plasma PPi levels.

### Determinants of circulating PPi levels; association with phosphate, ALP activity, BMI and coronary artery calcification

Hydrolysis of PPi by tissue-nonspecific ALP is a key regulatory mechanism of PPi homeostasis. This is evidenced by elevated plasma PPi in hypophosphatasia patients, deficient in ALP activity (50), and by studies demonstrating that ALP decreases plasma PPi concentration and prevents arterial calcification in CKD rodent models (51). Furthermore, physiological concentrations of Pi inhibit ALP enzymatic activity (52), suggesting reciprocal regulation.

Our results demonstrate a strong positive association between plasma PPi and serum Pi, and a robust inverse relationship between PPi and ALP activity across univariate and multivariate models. These findings are consistent with observations in CKD and end-stage renal disease (36, 53), and in patients with aortic valve calcification (38), establishing that ALP activity and Pi concentration are major determinants of plasma PPi levels in cardiovascular patients.

In addition, we identified two novel associations. Plasma PPi levels showed a consistent positive correlation with BMI, an association not previously reported. Patient-reported vitamin D intake also positively correlated with PPi levels. While experimental models link vitamin D to vascular calcification (54, 55), a direct relationship with PPi has not yet been documented in humans. However, given that vitamin D levels were not directly measured in our study, this finding warrants further investigation. Age, sex, and diabetes status were not significantly associated with plasma PPi in our adjusted models, although non-significant trends were observed. Females tended to have slightly higher PPi levels, while individuals with diabetes showed lower levels. This is in line with previously published results (37, 49, 56, 57); however, the influence of these demographic and clinical variables on PPi regulation remains uncertain and requires further investigation.

In univariate analyses, the inverse relationship between plasma PPi and CAC did not reach statistical significance. However, in multivariate models adjusting for traditional cardiovascular risk factors and potential confounders, left circumflex artery calcification (LCx CAC) emerged as a significant inverse predictor of PPi levels. Of note, a similar negative trend was observed for total coronary calcium burden, though it did not attain statistical significance. These findings suggest that the extent of coronary calcification may influence circulating PPi levels, with advancing disease stage potentially affecting systemic PPi concentrations.

### Determinants of coronary artery calcification; inverse association with plasma PPi and alkaline phosphatase activity

In multivariate models, plasma PPi emerged as an independent inverse correlate of left circumflex artery (LCx) calcification, consistent with PPi’s established role as an endogenous inhibitor of vascular mineralization. While this association reached statistical significance for LCx CAC, the corresponding association with sumCAC was directionally consistent but did not attain significance, likely reflecting reduced statistical power and increased heterogeneity associated with aggregated CAC measures. Although a segment-specific effect cannot be excluded, the present data do not allow distinction between true regional specificity and reduced sensitivity of global CAC measures in a cohort of this size. Accordingly, larger studies will be required to determine whether segment-specific associations persist in adequately powered cohorts.

In multivariate analyses, diabetes mellitus (DM) exhibited an inverse association with LCx coronary artery calcification, while HbA1c showed a positive association. This apparent paradox is likely multifactorial and reflects both clinical and methodological effects. A diagnosis of DM may serve as a proxy for intensified cardiovascular risk management, including glucose-lowering therapy, statin use, and lifestyle modification, which may attenuate CAC progression. In contrast, HbA1c represents a continuous measure of glycemic burden and cumulative metabolic stress, capturing subclinical dysglycemia and residual hyperglycemia despite treatment. Accordingly, HbA1c may more accurately reflect the pathophysiological contribution of glucose dysregulation to coronary calcification than a binary diabetes diagnosis. In addition, survivor and referral biases inherent to cardiovascular cohorts may contribute to this observation, as patients with long-standing diabetes and advanced coronary calcification may have experienced prior cardiovascular events and thus be underrepresented in the present cohort. Finally, diabetes is associated with medial vascular calcification, which may be less accurately captured by coronary artery calcium scoring that primarily reflects intimal calcification (13).

In addition to LCx CAC’s strong positive association with serum phosphate, coronary artery calcification was inversely associated with both PPi and alkaline phosphatase (ALP) activity in the calcifying patients. This unexpected finding suggest that systemic PPi availability and ALP activity may jointly reflect and influence local vascular mineralization processes. The directionally consistent but non-significant associations of sumCAC with PPi, ALP, and phosphate support preservation of these relationships at the global coronary level, albeit attenuated by increased heterogeneity and limited statistical power. Together, these clinical associations suggest the presence of a local compensatory PPi response to mineral stress in calcifying coronary arteries.

### Evidence from coronary artery explants and calcifying VSMC supports a local compensatory PPi response

*Ex-vivo* data in conditioned media from human coronary artery explants demonstrated significantly elevated PPi levels from atherosclerotic arteries compared to healthy, lesion-free vessels. These findings suggest that atherosclerotic or calcified vascular tissue may actively upregulate local PPi release. Supporting this observation, phosphate-induced calcification of A7r5 rat VSMCs also resulted in a time-dependent increase in extracellular PPi levels. The proposed mechanism—local vascular PPi release in response to calcific or atherosclerotic stress—may reflect an adaptive, compensatory response mitigating further mineral deposition in advanced disease. Notably, this concept introduces a new layer of PPi homeostasis under pathological conditions: while systemic PPi decline may promote disease progression, local PPi production may be enhanced to offset this deficiency. Further studies are necessary to validate this potentially clinically relevant compensatory mechanism and to define its role in coronary artery calcification, coronary artery disease, and broadly, in vascular calcification.

### Study limitations

The observations of the study should be interpreted in the context of the cross-sectional design and warrant validation in longitudinal cohorts. The present prospective study applied rigorous patient selection and stringent preanalytical and analytical procedures required for reliable plasma PPi quantification. Still, circadian variability in plasma PPi levels may have introduced residual confounding despite standardized sample collection protocols. Vitamin D intake was self-reported, and serum vitamin D levels were not directly measured. Although patients with overt kidney dysfunction or mineral metabolism disorders were excluded, individuals with diabetes mellitus and hypertension were included due to their high prevalence in the cardiovascular population and were adjusted for in the statistical analyses.

## Conclusion

This study provides the first clinical evidence linking circulating PPi levels to coronary artery calcification. Plasma PPi was inversely associated with left circumflex artery (LCx) calcification, and both PPi and ALP activity independently predicted LCx CAC in patients with established calcification. We also identified associations of PPi with serum phosphate, ALP, body mass index, and vitamin D intake. Ex vivo and in vitro analyses revealed increased PPi release from atherosclerotic coronary arteries and calcifying vascular smooth muscle cells, supporting a potential local compensatory mechanism. These findings suggest a dual model in which systemic PPi deficiency promotes vascular calcification, while diseased arteries may locally upregulate PPi to limit further mineral deposition. Validation in larger cohorts could establish PPi as a biomarker and therapeutic target in coronary artery disease.

## Supporting information

Supplementary Results

## NON-STANDARD ABBREVIATIONS AND ACRONYMS

ABCC6: ATP binding cassette subfamily C member 6
AICc: corrected Akaike information criterion
ALP: alkaline phosphatase
ANK(H): progressive ankylosis protein (homologue)
CAD: coronary artery disease
CAC: coronary artery calcium or coronary artery calcification
CCTA: cardiac computed tomography angiography
CT: computed tomography
DM: diabetes mellitus
eGFR: estimated glomerular filtration rate
ENPP1: ectonucleotide pyrophosphatase/phosphodiesterase 1
HbA1c: hemoglobin A1c
HT: hypertension
LDL-C: low density lipoprotein cholesterol
HDL-C: high density lipoprotein cholesterol
LAD: left anterior descending artery
LCx: left circumflex artery
LCx: CAC Agatston score of the left circumflex artery
LM: left main coronary artery
Lp(a): lipoprotein(a)
MLR: multiple linear regression
Pi: serum inorganic phosphate
Ppi: plasma inorganic pyrophosphate
PXE: pseudoxanthoma elasticum
RCA: right coronary artery
sumCAC: Agatston score of the total coronary artery calcium
TAI: thrombocyte aggregation inhibitor
TG: triglycerides
TNAP: tissue non-specific alkaline phosphatase
VSMC: vascular smooth muscle cell

## Data Availability

All data produced in the present study are available upon reasonable request to the authors

## ACKNOWLEDGEMENT

We thank Zsófia Demjén Nagy and Áron Nagy for their assistance in the preparation of patient samples. The graphical abstract contains illustrations provided by Servier Medical Art (https://smart.servier.com/), licensed under CC BY 4.0 (https://creativecommons.org/licenses/by/4.0/).

## TABLES

**Table 1.**
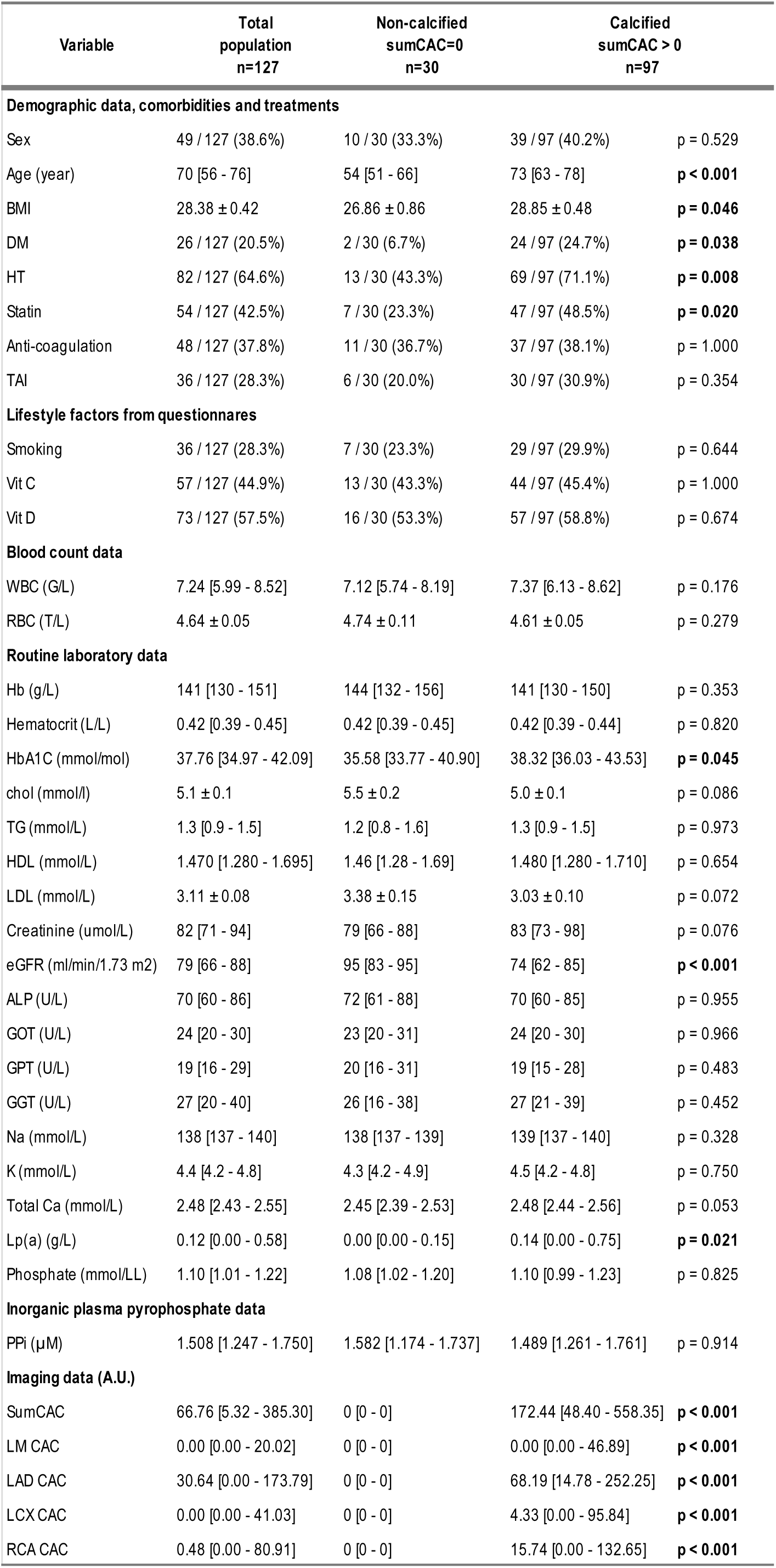
Patient characteristics and stratification. Demographic, clinical, self-reported and routine laboratory data of patients with plasma PPi concentration and coronary artery calcification (CAC) determined by the Agatston method. Based on CAC scores, patients are classified as non-calcified (sumCAC = 0; n = 30) or calcified (sumCAC > 0; n = 97) group. Statistical significance (p-value < 0.05) compared to values of non-calcified patients is indicated in bold. ALP, serum alkaline phosphatase activity; A.U., arbitrary unit; BMI, body mass index; chol, total cholesterol; DM, diabetes mellitus; eGFR, estimated glomerular filtration rate; GGT, gamma-glutamyl transferase; GOT, glutamic-oxaloacetic transaminase; GPT (U/L), glutamic pyruvic transaminase; Hb, hemoglobin; HbA1c, hemoglobin A1c; HDL-C, high density lipoprotein cholesterol; HT, hypertension; K, potassium; LAD CAC, Agatston score of the left anterior descending artery; LCx CAC, Agatston score of the left circumflex artery; LDL-C, low density lipoprotein cholesterol; LM CAC, Agatston score of the left main coronary artery; Lp(a), lipoprotein(a); Na, sodium; Pi, serum inorganic phosphate; PPi, plasma inorganic pyrophosphate; RBC, red blood cell; RCA CAC, Agatston score of the right coronary artery; Sum CAC, Agatston score of the total coronary artery calcium; TAI, thrombocyte aggregation inhibitor; TG, triglycerides; Total Ca, total calcium; vit C, vitamin C; vit D, vitamin D; WBC, white blood cell.

| Variable | Total population<br>n=127 | Non-calcified<br>sumCAC=0<br>n=30 | Calcified<br>sumCAC > 0<br>n=97 |  |
| --- | --- | --- | --- | --- |
| **Demographic data, comorbidities and treatments** |  |  |  |  |
| Sex | 49 / 127 (38.6%) | 10 / 30 (33.3%) | 39 / 97 (40.2%) | p = 0.529 |
| Age (year) | 70 [56 - 76] | 54 [51 - 66] | 73 [63 - 78] | p < 0.001 |
| BMI | 28.38 ± 0.42 | 26.86 ± 0.86 | 28.85 ± 0.48 | p = 0.046 |
| DM | 26 / 127 (20.5%) | 2 / 30 (6.7%) | 24 / 97 (24.7%) | p = 0.038 |
| HT | 82 / 127 (64.6%) | 13 / 30 (43.3%) | 69 / 97 (71.1%) | p = 0.008 |
| Statin | 54 / 127 (42.5%) | 7 / 30 (23.3%) | 47 / 97 (48.5%) | p = 0.020 |
| Anti-coagulation | 48 / 127 (37.8%) | 11 / 30 (36.7%) | 37 / 97 (38.1%) | p = 1.000 |
| TAI | 36 / 127 (28.3%) | 6 / 30 (20.0%) | 30 / 97 (30.9%) | p = 0.354 |
| **Lifestyle factors from questionnaires** |  |  |  |  |
| Smoking | 36 / 127 (28.3%) | 7 / 30 (23.3%) | 29 / 97 (29.9%) | p = 0.644 |
| Vit C | 57 / 127 (44.9%) | 13 / 30 (43.3%) | 44 / 97 (45.4%) | p = 1.000 |
| Vit D | 73 / 127 (57.5%) | 16 / 30 (53.3%) | 57 / 97 (58.8%) | p = 0.674 |
| **Blood count data** |  |  |  |  |
| WBC (G/L) | 7.24 [5.99 - 8.52] | 7.12 [5.74 - 8.19] | 7.37 [6.13 - 8.62] | p = 0.176 |
| RBC (T/L) | 4.64 ± 0.05 | 4.74 ± 0.11 | 4.61 ± 0.05 | p = 0.279 |
| **Routine laboratory data** |  |  |  |  |
| Hb (g/L) | 141 [130 - 151] | 144 [132 - 156] | 141 [130 - 150] | p = 0.353 |
| Hematocrit (L/L) | 0.42 [0.39 - 0.45] | 0.42 [0.39 - 0.45] | 0.42 [0.39 - 0.44] | p = 0.820 |
| HbA1C (mmol/mol) | 37.76 [34.97 - 42.09] | 35.58 [33.77 - 40.90] | 38.32 [36.03 - 43.53] | p = 0.045 |
| chol (mmol/l) | 5.1 ± 0.1 | 5.5 ± 0.2 | 5.0 ± 0.1 | p = 0.086 |
| TG (mmol/L) | 1.3 [0.9 - 1.5] | 1.2 [0.8 - 1.6] | 1.3 [0.9 - 1.5] | p = 0.973 |
| HDL (mmol/L) | 1.470 [1.280 - 1.695] | 1.46 [1.28 - 1.69] | 1.480 [1.280 - 1.710] | p = 0.654 |
| LDL (mmol/L) | 3.11 ± 0.08 | 3.38 ± 0.15 | 3.03 ± 0.10 | p = 0.072 |
| Creatinine (umol/L) | 82 [71 - 94] | 79 [66 - 88] | 83 [73 - 98] | p = 0.076 |
| eGFR (ml/min/1.73 m2) | 79 [66 - 88] | 95 [83 - 95] | 74 [62 - 85] | p < 0.001 |
| ALP (U/L) | 70 [60 - 86] | 72 [61 - 88] | 70 [60 - 85] | p = 0.955 |
| GOT (U/L) | 24 [20 - 30] | 23 [20 - 31] | 24 [20 - 30] | p = 0.966 |
| GPT (U/L) | 19 [16 - 29] | 20 [16 - 31] | 19 [15 - 28] | p = 0.483 |
| GGT (U/L) | 27 [20 - 40] | 26 [16 - 38] | 27 [21 - 39] | p = 0.452 |
| Na (mmol/L) | 138 [137 - 140] | 138 [137 - 139] | 139 [137 - 140] | p = 0.328 |
| K (mmol/L) | 4.4 [4.2 - 4.8] | 4.3 [4.2 - 4.9] | 4.5 [4.2 - 4.8] | p = 0.750 |
| Total Ca (mmol/L) | 2.48 [2.43 - 2.55] | 2.45 [2.39 - 2.53] | 2.48 [2.44 - 2.56] | p = 0.053 |
| Lp(a) (g/L) | 0.12 [0.00 - 0.58] | 0.00 [0.00 - 0.15] | 0.14 [0.00 - 0.75] | p = 0.021 |
| Phosphate (mmol/LL) | 1.10 [1.01 - 1.22] | 1.08 [1.02 - 1.20] | 1.10 [0.99 - 1.23] | p = 0.825 |
| **Inorganic plasma pyrophosphate data** |  |  |  |  |
| PPi (μM) | 1.508 [1.247 - 1.750] | 1.582 [1.174 - 1.737] | 1.489 [1.261 - 1.761] | p = 0.914 |
| **Imaging data (A.U.)** |  |  |  |  |
| SumCAC | 66.76 [5.32 - 385.30] | 0 [0 - 0] | 172.44 [48.40 - 558.35] | p < 0.001 |
| LM CAC | 0.00 [0.00 - 20.02] | 0 [0 - 0] | 0.00 [0.00 - 46.89] | p < 0.001 |
| LAD CAC | 30.64 [0.00 - 173.79] | 0 [0 - 0] | 68.19 [14.78 - 252.25] | p < 0.001 |
| LCX CAC | 0.00 [0.00 - 41.03] | 0 [0 - 0] | 4.33 [0.00 - 95.84] | p < 0.001 |
| RCA CAC | 0.48 [0.00 - 80.91] | 0 [0 - 0] | 15.74 [0.00 - 132.65] | p < 0.001 |

