## Supplementary Results for "Inverse association between coronary artery calcification and plasma pyrophosphate: systemic deficiency and local compensation"

### SUPPLEMENTARY FIGURES

Supplementary Figure 1

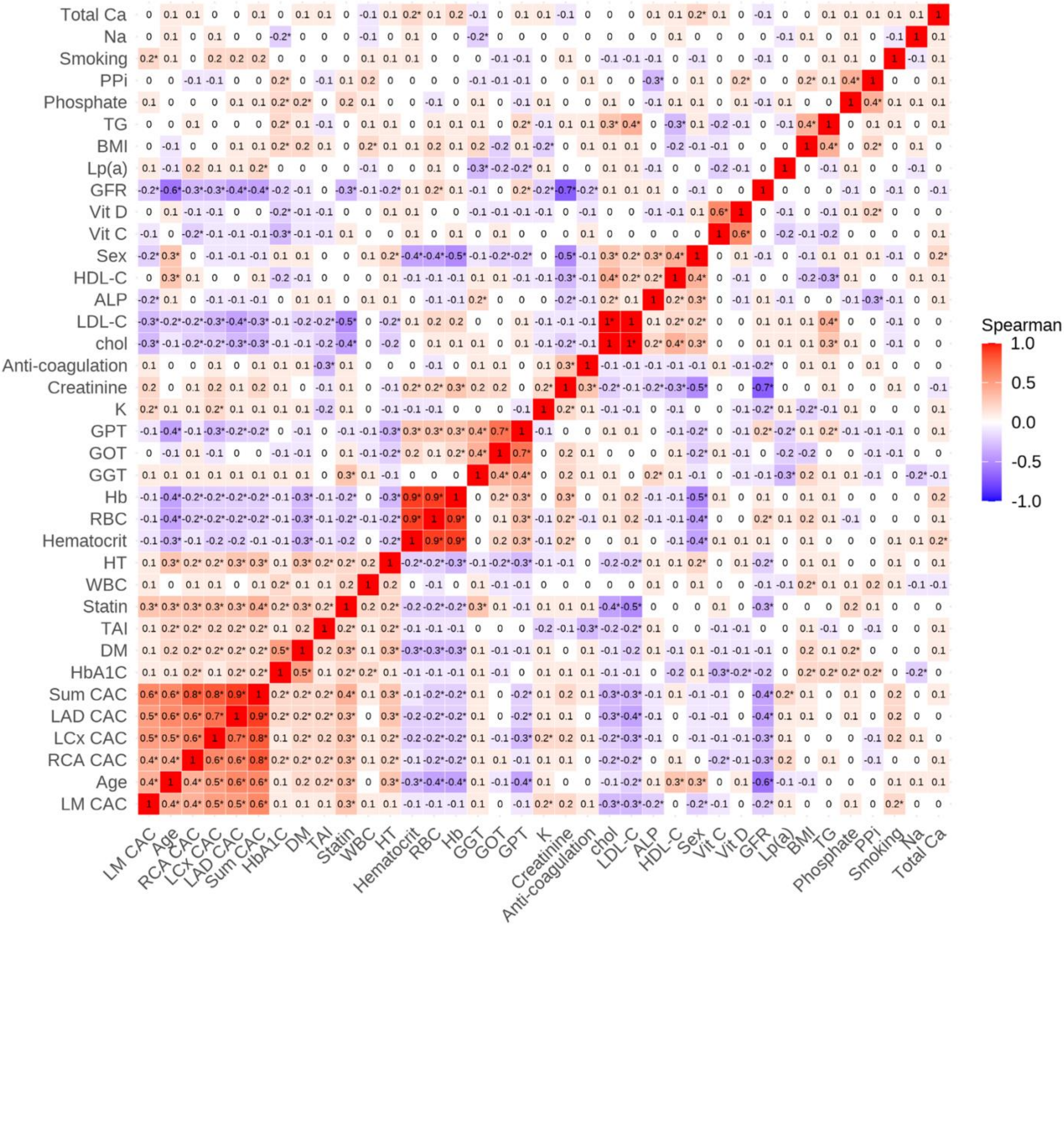

Supplementary Figure 1

**Spearman correlation of demographic, clinical and biochemical variables.** Heatmap illustrating Spearman rank correlation with coefficients (ρ) between continuous and categorical variables. Statistically significant correlations are indicated by asterisks. Positive correlations are shown in red and negative correlations in blue, with intensity proportional to the strength of the association. ALP, serum alkaline phosphatase activity; BMI, body mass index; chol, total cholesterol; DM, diabetes mellitus; eGFR, estimated glomerular filtration rate; GGT, gamma-glutamyl transferase; GOT, glutamic-oxaloacetic transaminase; GPT (U/L), glutamic pyruvic transaminase; Hb, hemoglobin; HbA1c, hemoglobin A1c; HDL-C, high density lipoprotein cholesterol; HT, hypertension; K, potassium; LAD CAC, Agatston score of the left anterior descending artery; LCx CAC, Agatston score of the left circumflex artery; LDL-C, low density lipoprotein cholesterol; LM CAC, Agatston score of the left main coronary artery; Lp(a), lipoprotein(a); Na, sodium; PPi, plasma inorganic pyrophosphate; RBC, red blood cell; RCA CAC, Agatston score of the right coronary artery; Sum CAC, Agatston score of the total coronary artery calcium; TAI, thrombocyte aggregation inhibitor; TG, triglycerides; Total Ca, total calcium; vit C, vitamin C; vit D, vitamin D; WBC, white blood cell.

Supplementary Figure 2

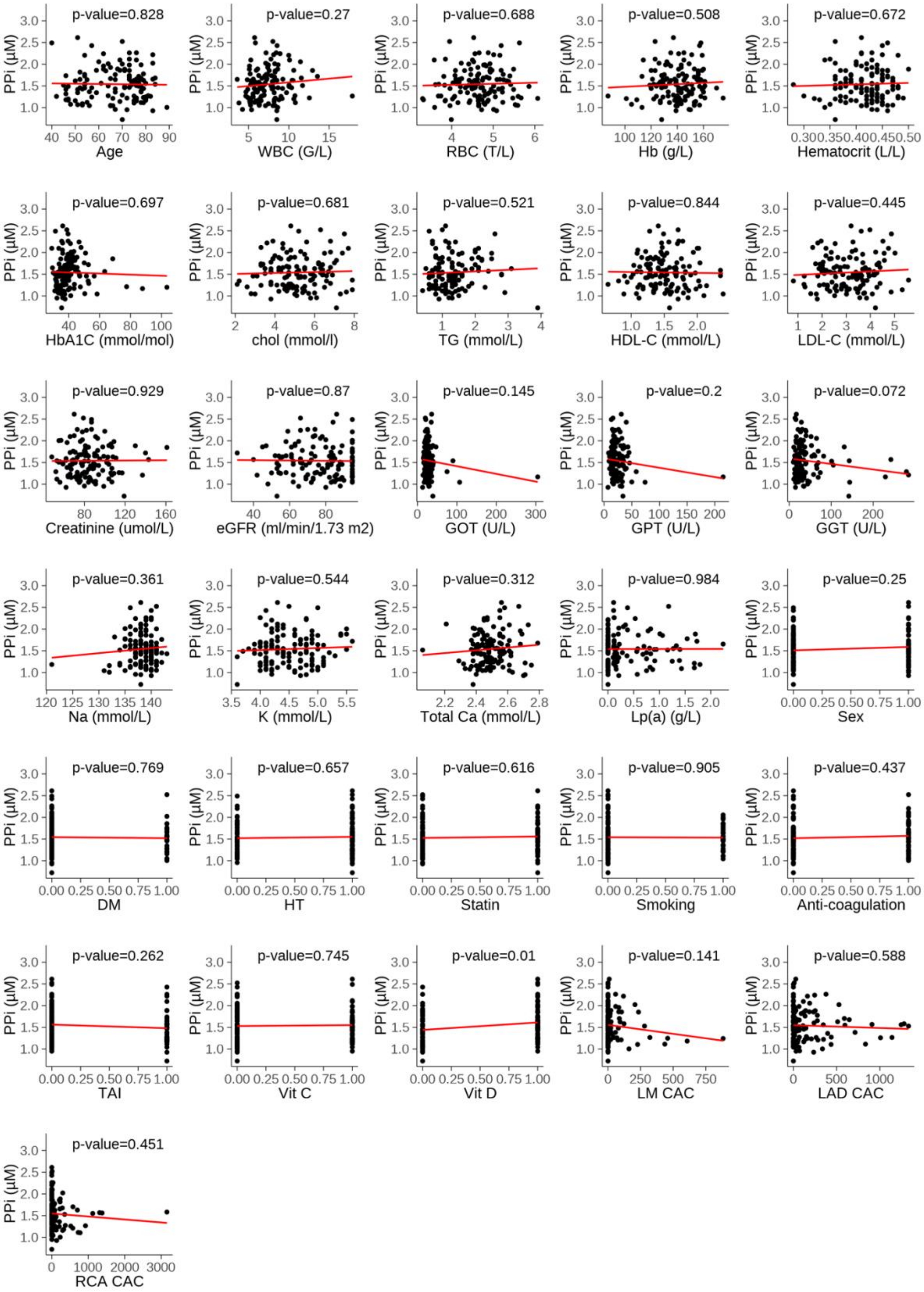

Supplementary Figure 2

**Univariate associations with plasma PPi.** Univariate associations between plasma pyrophosphate (PPi) levels and demographic, clinical or biochemical variables. Scatter plots with linear regression lines, fitting parameters and the significance of the correlations. Univariate plots depicting PPi levels in the function of ALP, Pi, BMI, LCX CAC and sumCAC are shown in the Main Figures (Figure 1C). ALP, serum alkaline phosphatase activity; BMI, body mass index; chol, total cholesterol; DM, diabetes mellitus; eGFR, estimated glomerular filtration rate; GGT, gamma-glutamyl transferase; GOT, glutamic-oxaloacetic transaminase; GPT (U/L), glutamic pyruvic transaminase; Hb, hemoglobin; HbA1c, hemoglobin A1c; HDL-C, high density lipoprotein cholesterol; HT, hypertension; K, potassium; LAD CAC, Agatston score of the left anterior descending artery; LCx CAC, Agatston score of the left circumflex artery; LDL-C, low density lipoprotein cholesterol; LM CAC, Agatston score of the left main coronary artery; Lp(a), lipoprotein(a); Na, sodium; Pi, serum inorganic phosphate; PPi, plasma inorganic pyrophosphate; RBC, red blood cell; RCA CAC, Agatston score of the right coronary artery; Sum CAC, Agatston score of the total coronary artery calcium; TAI, thrombocyte aggregation inhibitor; TG, triglycerides; Total Ca, total calcium; vit C, vitamin C; vit D, vitamin D; WBC, white blood cell.

Supplementary Figure 3

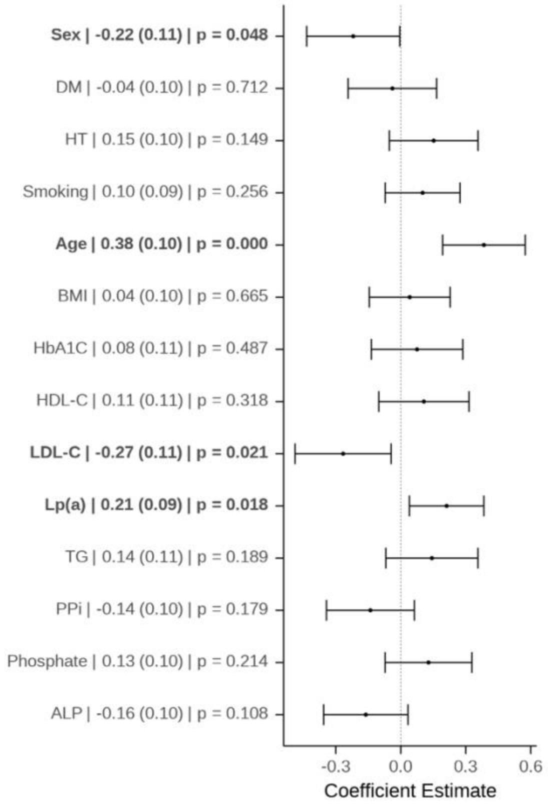


Supplementary Figure 3

**Determinants of total calcification burden in multiple linear regression (MLR).** Determinants for association with sumCAC score in MLR with adjustment for major confounders, effect size, SE and p value are indicated. In calcified individuals, sumCAC scores were significantly associated with male sex, age, LDL-C, and lipoprotein(a) levels, while non-significant inverse trends were observed between sumCAC and plasma PPi as well as ALP activity, whereas a non-significant positive trend was noted with serum Pi levels.

ALP, serum alkaline phosphatase activity; BMI, body mass index; DM, diabetes mellitus; HbA1c, hemoglobin A1c; HDL-C, high density lipoprotein cholesterol; HT, hypertension; LDL-C, low density lipoprotein cholesterol; Lp(a), lipoprotein(a); PPi, plasma inorganic pyrophosphate; TG, triglycerides.
